# Strong Effect of Demographic Changes on Tuberculosis Susceptibility in South Africa

**DOI:** 10.1101/2023.11.02.23297990

**Authors:** Oshiomah P. Oyageshio, Justin W. Myrick, Jamie Saayman, Lena van der Westhuizen, Dana Al-Hindi, Austin W. Reynolds, Noah Zaitlen, Caitlin Uren, Marlo Möller, Brenna M. Henn

## Abstract

South Africa is among the world’s top eight TB burden countries, and despite a focus on HIV-TB co-infection, most of the population living with TB are not HIV co-infected. The disease is endemic across the country with 80-90% exposure by adulthood. We investigated epidemiological risk factors for tuberculosis (TB) in the Northern Cape Province, South Africa: an understudied TB endemic region with extreme TB incidence (645/100,000) and the lowest provincial population density. We leveraged the population’s high TB incidence and community transmission to design a case-control study with population-based controls, reflecting similar mechanisms of exposure between the groups. We recruited 1,126 participants with suspected TB from 12 community health clinics, and generated a cohort of 878 individuals (cases =374, controls =504) after implementing our enrollment criteria. All participants were GeneXpert Ultra tested for active TB by a local clinic. We assessed important risk factors for active TB using logistic regression and random forest modeling. Additionally, a subset of individuals were genotyped to determine genome-wide ancestry components. Male gender had the strongest effect on TB risk (OR: 2.87 [95% CI: 2.1-3.8]); smoking and alcohol consumption did not significantly increase TB risk. We identified two interactions: age by socioeconomic status (SES) and birthplace by residence locality on TB risk (OR = 3.05, p = 0.016) – where rural birthplace but town residence was the highest risk category. Finally, participants had a majority Khoe-San ancestry, typically greater than 50%. Epidemiological risk factors for this cohort differ from other global populations. The significant interaction effects reflect rapid changes in SES and mobility over recent generations and strongly impact TB risk in the Northern Cape of South Africa. Our models show that such risk factors combined explain 16% of the variance (r^2^) in case/control status.

## Introduction

Tuberculosis (TB) is among the world’s leading causes of death due to infectious disease, recently surpassed by COVID-19 (1). The TB causative agent, *Mycobacterium tuberculosis (M.tb),* is an obligate, exclusive *Homo sapiens* pathogen mainly infecting the lungs, and sometimes other organs (2,3). Determinants of active TB progression are multifaceted including human host genetics, nutrition, social and economic conditions, behavior, and sex-specific biology (1,4,5). The extent of these determinants’ effects varies across and within populations, necessitating epidemiological studies in differing contexts and communities (5). These factors have also been shown to vary between low and high/intermediate-incidence populations, with lower odds ratios in high/intermediate-incidence populations (6). Here, we characterize the TB epidemiology of a district in the Northern Cape Province, South Africa, a TB-endemic region with relatively low HIV.

South Africa is amongst the top 30 ‘high burden’ countries, burdened by TB, TB/HIV co-infection, and multi-drug resistance or rifampicin-resistant TB (MDR/RR-TB). TB is South Africa’s leading natural cause of death (7) with an extremely high prevalence (852/100,000, (8)) and accounts for 3.3% of all global TB cases (1). HIV is commonly identified as the leading risk factor for TB. In South Africa, 59% of TB patients on a TB programme (screened by a clinician and on TB medication) are co-infected with HIV. South Africa’s first national TB prevalence survey (n=35,000), however, found only 28% of TB cases were co-infected with HIV(8). This discrepancy is partly explained by those with TB who go undetected, mainly symptomatic men not living with HIV who have limited clinical contact (8). Many individuals who are diagnostically TB+ may also go undetected because 78% of TB+ HIV-individuals exhibit only one or no classic TB symptoms (e.g. cough for two weeks, fever, night sweats, and weight loss; 61% have no symptoms)(8).

In case-control studies, controls should have similar disease exposure profiles to the cases. Population-based controls risk differential disease exposure, a concern in low-incidence populations that can bias statistical associations. However, in high-incidence populations with well-characterized disease burdens and transmission, population controls greatly improve statistical power (9). In South Africa, TB is community spread moreover than household (10,11) and TB latency increases with age, reaching 80% by age 30 (12–15), an epidemiological scenario that ensures cases and controls have approximate disease exposure profiles.

*Mycobacterium tuberculosis* has a long coevolutionary history with different human populations likely leading to population-specific genetic signatures (2,16). TB susceptibility phenotypes have a heritability of 11-92% (17), yet few critical genetic variants have replicated across genome-wide association studies (GWAS) (18), potentially reflecting these population-specific signatures. This result has spurred several studies to examine the relationship between genetic ancestry and TB risk (19–23). For instance, native Amerindian ancestry was shown to be a risk factor for TB progression in an admixed Amazonian population, and genetic variants in Peruvian populations have been associated with early active TB progression (19–21). A major challenge in identifying genetic risk factors associated with TB progression is decoupling the social and environmental effects that accompany ancestry. To this end, Asgari et al. controlled for environmental effects (e.g., sanitation, water supply, and socioeconomic status [SES])) and found indigenous Peruvian ancestry to be an independent, significant predictor of TB progression. Chimusa et al. also corrected for SES and demonstrated an association between Khoe-San ancestry and TB progression in South Africa (22).

In this study, we investigated ancestry proportions as well as several common TB epidemiological variables identified in earlier studies (24). Smoking, alcohol consumption, and intravenous drug use have independently been associated with TB. Meta-analyses have found alcohol use and smoking (25,26), and specifically heavy alcohol use (26–28) to increase TB risk, though not always consistently. Our study is part of the Northern Cape Tuberculosis Project (NCTB), investigating the human-host genetics of TB among admixed Khoe-San descent populations in rural or peri-urban communities. Characterizing the TB epidemiology in this region will identify nongenetic risk factors that can serve as control variables in future genetic studies of TB risk.

## Methods

### Research ethics statement

This study has been approved by the Health Research Ethics Committee (HREC) of Stellenbosch University (N11/07/210A) and the Northern Cape Department of Health (NC2015/008). All participants were adults (18 years and older) and provided written informed formal consent. Authors Justin W. Myrick, Jamie Saayman, Lena van der Westhuizen and Marlo Möller had access to identifiable information about participants as they were directly involved in data collection or database management. Access to these records commenced on <u>26th January</u> <u>2016,</u> and is still ongoing as it is an integral part of the ongoing Northern Cape Tuberculosis Project (NCTB).

### Study Design and Recruitment

Participants (18 years and older) provided written informed consent and were recruited from 12 community health clinics from the ZF Mgcawu district in the Northern Cape Province of South Africa from <u>26th January 2016 - 15 May 2017, and 11 December 2018 - 11 March 2020</u>. Community health clinics are the front line for TB screening and treatment, visited by 87% of people who seek TB care (8). TB nurses referred patients with suspected TB (with ≥2 TB symptoms: cough for ≥2 weeks, night sweats, weight loss, and fever ≥2 weeks or a TB contact) and TB patients to our on-site RAs. All study participants took a clinic-administered sputum GeneXpert Ultra test for active TB at the time of the study interview and provided saliva for genotyping. Clinic medical charts were accessed by a staff research nurse to record GeneXpert test results and verify HIV status and TB history.

### Case-Control Assignment

Cases and controls were assigned reckoning the participant’s medical charts and self-reported data (see Fig. 1). Cases include anyone with active pulmonary TB in their lifetime *and* are HIV-negative and followed two tracks: 1) Clinically confirmed active TB (n= 343) and 2) self-reported past TB episode(s) (n=228). GeneXpert results, diagnostic test date, TB strain (drug resistance), and TB medication regimen were used to determine clinically confirmed progression to active TB. Past TB episodes are self-reported, mainly due to older medical charts not reliably available, discarded, or difficult to locate by clinic staff.

**Fig 1.**
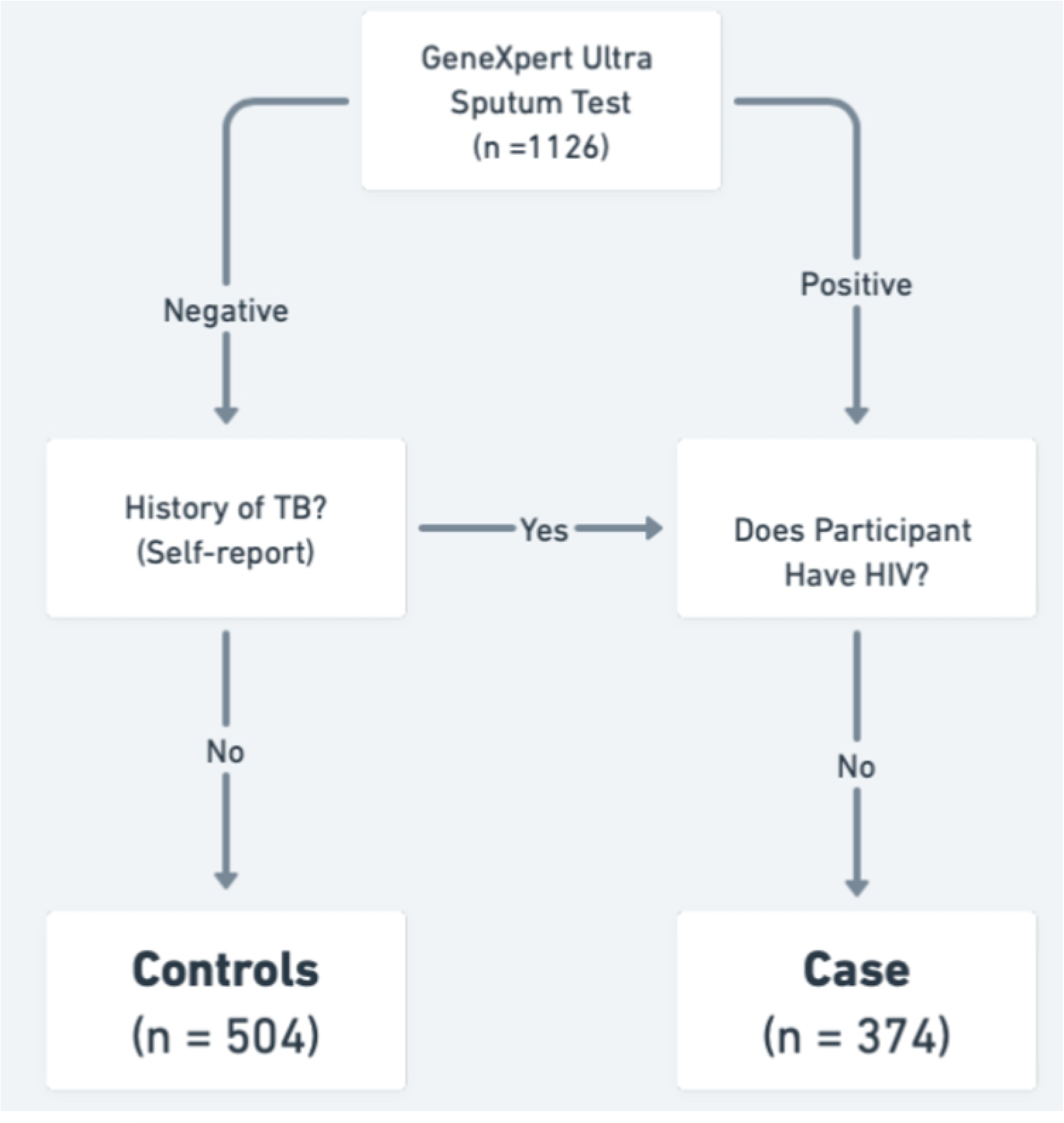
Case-Control Decision Tree. Study participants were categorized as cases or controls based on medical record information and self-reported data. All participants were GeneXpert tested for active TB infection at the time of enrollment. Past TB episodes were self-reported and cross-referenced with medical records when available.

Controls are patients with suspected TB who have a negative GeneXpert Ultra result and have no history of active pulmonary TB at the time of study enrollment and are largely assumed to be latently infected with *M.tb* (LTBI). A majority of the population in high TB burden South African suburbs are LTBI, 88% by ages 31-35 (12,13) and studies have consistently shown LTBI in South Africa to be above 75% by age 25, increasing across adulthood (14). Our population-control design relies on population-wide TB exposure, as traditional screening methods, tuberculin skin test (TST) and interferon-gamma release assay (IGRA; e.g., QuantiFERON), are limited both in the concordance and positive predictive value (29,30). IGRA and TST are used to infer *M.tb* infection, but cannot be used to determine previous exposure to the bacterium. Certain individuals living in high *M.tb* exposed populations test persistently negative for these tests and do not develop active disease, but display *Mtb-*specific antibody titres. These individuals are known as “resisters” or “early clearers” (31,32).

Our exclusion criteria removed participants with unknown TB or HIV status, as well as individuals with dual HIV and TB infections.

### Study covariates

We collected demographic information that included date of birth, place of birth, current residence, self-identified gender, self-reported ethnic identity, and parental ethnic identities.

Behavioral variables include smoking and alcohol consumption (See Supplementary Materials in S1 Text). In our analyses, we only used binary measures for smoking and alcohol (“Do you smoke?”, “Do you drink alcohol?”). Residence and birthplace locations are categorized as rural (≤2000 people) and town (>2,000 people). Population size was derived from the South African census and when census data was absent, e.g., a farm, we used Google Earth (earth.google.com) to estimate population size based on the number of dwellings. Age was used as a continuous variable for all analyses and binned for calculating empirical odds (see Fig 2B). SES was operationalized as someone’s number of years of education, i.e., the highest completed level of education. McKenzie et al. have shown education level, in this dataset, positively predicts body mass index in TB controls, tracking access to resources and food security (33).

**Fig 2.**
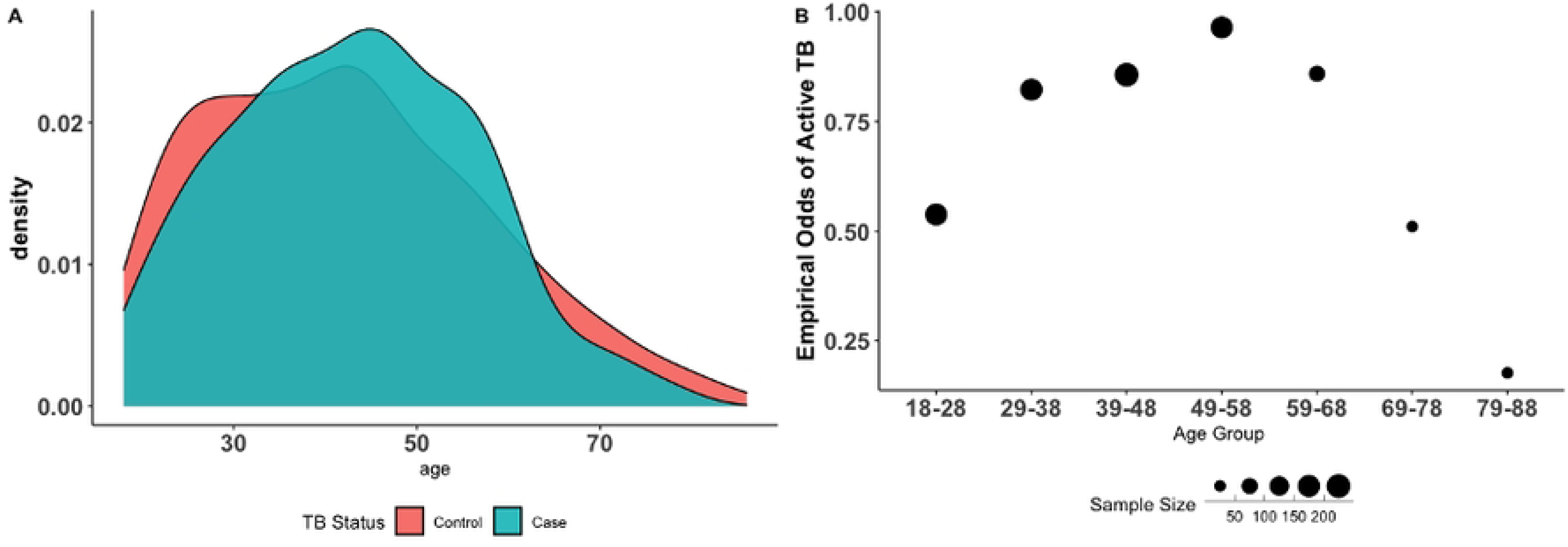
Case-Control status shifts across Age groups. A) Overlapping density plots of age distribution stratified by TB status (n= 878). At the oldest and youngest ages, most of our study participants are cases whilst at middle-age groups, the majority are controls. B) Empirical odds of active TB by age group. The x-axis bins our participants into 7 age groups and the y-axis: the empirical odds of active TB. Empirical odds are calculated by dividing the number of controls divided by the number of cases in each age bin. The size of the dots corresponds to the sample size of the age group. Our data reveal a signal of survivor bias. Since age is a cumulative measure of exposure, the empirical odds of TB should increase with age. This pattern is observed from our youngest age group up to age 58. The empirical odds of TB progressively decrease after age 58. Older age groups are biased towards controls due to the mortality of TB.

### Data Analyses

Statistical analyses were performed in R (version 4.0.2). We calculated Pearson correlations with the R package *ggcorrplot*. All categorical variables were numerically coded to “0” and “1”. Classification models for our binary, qualitative dependent variable (“case”/ “control”) included logistic regression and random forest—a machine learning classifier robust to non-linear associations and unknown variable interactions (34) (see Supplementary Materials in S1 Text). Random forest is a growing analytic tool in epidemiology (35–37). The coefficients of the logistic regression models were converted to odds ratios using the R package *gtools* (38), and marginal effects were plotted using the R package *effects* (39). Each model was Bonferroni corrected by dividing, 0.05, by the number of variables in said model.

Our first model, the common risk factor model (n=878), includes seven covariates known to be common risk factors for TB.

TB Status ∼ gender + smoking + drinking + diabetes + residence + age + SES Health disparities are one of the many consequences of apartheid in South Africa (40,41).

The end of apartheid improved social mobility and educational access, however, health disparities in the Northern Cape still remain (42). To capture the effect of lived experience vis-à-vis Apartheid on TB outcomes we designed the “SES model” (n=878). This model includes the common risk factor model and interacts with age and SES. Age is kept as a continuous variable because Apartheid was not a historically binary event.

TB Status ∼ common risk factor model + age * SES

Residing in an urban or rural environment is an established risk factor for TB status. In the “residence model”, we test the relationship between current residence and birthplace residence on TB status. Here, we build on the common risk factor model to include an interaction between current residence and birthplace. Setting this interaction allows us to examine four patterns, namely: rural birthplace to urban residence, urban birthplace to rural residence, lifetime rural residence, and lifetime urban residence.

TB Status ∼ common risk factor model + residence * birthplace

### Genetic Data Processing & Ancestry Estimation

Genetic data processing involved DNA extraction from saliva samples, genotyping for >2 million SNPs, common variant calling with GenomeStudio, rare variant calling with zCall, and further data cleaning using plink2 with specific parameters (Supplementary methods in S1 Text).Prior to genetic ancestry estimation, SNPs out of Hardy-Weinberg equilibrium (--hwe 0.001) and rare alleles (--maf 0.01) were removed from the dataset. The dataset was also pruned for linkage disequilibrium (--indep-pairwise 200 25 0.4). Individuals from Luhya, Maasai, Himba, British, Palestinian, Chinese, Bangladeshi, Tamil, Ju|’hoansi San, Khomani San, Nama populations were used as reference groups. Global ancestry estimates were calculated using ADMIXTURE v1.13(43). This was done in groups of maximally unrelated individuals to avoid biasing the ancestry estimates. ADMIXTURE was run for k=5 on unsupervised mode for each of the running groups. After matching clusters, we merged ancestry estimates across all running groups, averaging individuals that appeared in multiple running groups using pong (44)

## Results

### TB case-control classification

1,126 participants were partitioned into preliminary cases, preliminary controls, and unverified TB status (571,504, and 51 respectively; Table C in S1 Text). After excluding, participants with unverified TB status, preliminary cases with unverified HIV status, and participants co-infected with TB and HIV, 878 participants remained in the study (374 cases and 504 controls; Table A in S1 Text).

### Socio-behavioral covariates and demographics

Men and women were equally represented in the dataset (422:441, respectively, Table A in S1 Text). Men were more likely to drink alcohol (r = −0.14, p < 0.05; Fig I in S1 Text) and smoke (r = −0.22, p < 0.05; Fig I in S1 Text). Most of our participants smoked (66%) and 45% drank alcohol; smoking and drinking were moderately correlated with each other (*r* = 0.36, p < 0.05; Fig I in S1 Text). Women were more likely to have diabetes (r = 0.12, p < 0.05; Fig I in S1 Text) and, on average, had more education than men (female mean= 8.3 years, male mean = 7.7 years).

Cases and controls had similar distributions for age (mean = 43.1, SD =13.2 and mean =42.4, SD =15.2, respectively, Table A in S1 Text). “Age” is defined here as the age at the time of study enrollment and importantly, is a cumulative outcome: that is, it includes cases who currently and/or previously had TB, not the age of the TB episode. Age also captures the amount of time someone is exposed to TB. The empirical odds of active TB in our data reveal a signature of survivorship bias (Fig. 2B). We use the number of years of education as a proxy for “SES”.

The mean educational attainment is 8 years, equivalent to completing primary school, and similar between rural areas and towns (ANOVA, p > 0.1). In the ZF Mgcawu District census (45) 13% of people have not completed primary school compared to 25.3% of our participants. Age was moderately correlated with SES (r = −0.5, p < 0.05; Fig I in S1 Text) such that older participants tended to have lower SES.

### Ethnicity and Khoe-San Ancestry

Genetic ancestry analyses were performed for 159 participants (see Supplementary Methods in S1 Text) from the Northern Cape Tuberculosis Project on host-genetic susceptibility to TB. To our knowledge, this is the first study to report ancestry proportions of a clinical population in the Northern Cape Province, South Africa. Khoe-San ancestry varied across clinic locations (Fig. 3A) but remained the majority ancestry at each site (mean = 56%), followed by Bantu-speaking African ancestry (mean = 21%), European ancestry (mean = 16%), South Asian ancestry (mean = 5%), and East Asian ancestry (mean = 2%) (Fig. 3B).

**Fig 3.**
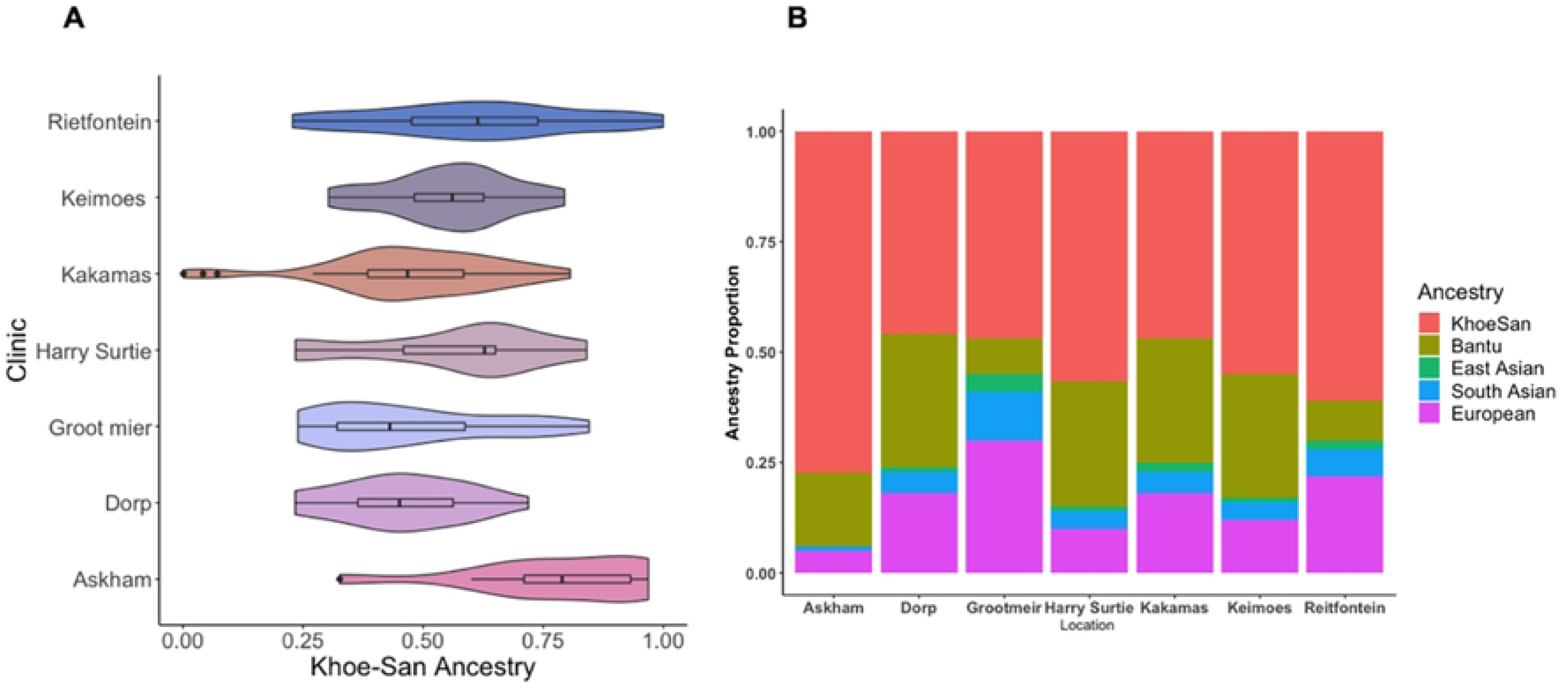
Khoe-San Ancestry is the Primary Genetic Ancestry in Clinics from the Northern Cape, South Africa. A subset of participants (n=159) was genotyped for preliminary ancestry analysis. A) The study population is admixed with 5 distinct ancestries with the Southern African indigenous Khoe-San ancestry being the largest proportion of ancestry across all study sites. (B) Although Khoe-San ancestry is the largest proportion of ancestry in our sample, it varies significantly across study sites.

Individuals were asked to self-identify their ethnicity without prompting. 88.4% of participants (both TB cases and controls) self-identify as, coloured, followed by 4.2% as a Khoe-San ethnicity (e.g., Nama, San), 4.6 % as Tswana, 1.3 % as Xhosa, and 1.9 % as “other”. Whilst we acknowledge that in some contexts the term, coloured, has derogatory connotations, it is a recognized ethnicity and used culturally in South Africa. People who self-identify using this term have different ancestries of different geographic origins, including the indigenous Khoe-San groups (e.g., Khoekhoe, San), Bantu-speaking, European, Indian, Malaysian (Southeast Asian) slaves, or people of mixed ancestry and their descendants (46).

### Logistic Regression Results

We designed three logistic regression models (47) to examine the risk factors’ odds ratios for the binary dependent variable, TB case/control. The common risk factor model included age, SES, gender, residence, smoking, diabetes, and alcohol as covariates. The SES model extended the common risk factor model to include an interaction between age and SES. Lastly, the residence model extended the common risk factor model to include an interaction between birthplace and current residence. The SES model (AIC = 1099; pseudo r^2^ = 17%, Table 1) performed slightly better than the common risk factor model (AIC= 1108; pseudo r^2^ =16%, Table 1). The residence model had a similar pseudo r^2^ (15% (Table B in S1 Text)) as the other two, however, we could not compare their AICs due to different sample sizes. All significance levels were Bonferroni corrected. This was carried out by dividing 0.05 by the number of variables used in the model.

**Table 1:** Odds ratios and p-values for the Demographic and Socio Behavioral Variables used in the Common Risk Factor Model and SES Model.

|  | Common Risk Factor Model |  | SES Model |  |
| --- | --- | --- | --- | --- |
|  | Odds Ratio (CI) | p-value | Odds Ratio (CI) | p-value |
| **Intercept** | 0.305 [0.13, 0.72] | 0.007 | 2.237 [0.46, 11.18] | 0.32 |
| **Gender -** | 2.87 [2.10, | 7.842e-12 | 2.85 [2.12, | 6.545e- |
| **Male** | 3.80] |  | 3.85] | 12 |
| **Years of Education (SES)** | 0.95 [0.90, 1.00] | 0.03 | 0.75 [0.63, 0.88] | 0.0007 |
| **Age** | 0.996 [0.99, 1.00] | 0.55 | 0.959 [0.931, 0.986] | 0.003 |
| **Drinks Alcohol - Yes** | 1.05 [0.77, 1.43] | 0.77 | 1.02 [0.75, 1.40] | 0.88 |
| **Diabetes - Yes** | 1.36 [0.73, 2.50] | 0.35 | 1.37 [0.74, 2.52] | 0.31 |
| **Current Residence - Rural (reference)** | 1 |  | 1 |  |
| **Current Residence - Town** | 2.88 [2.07, 4.02] | 5.316e-10 | 2.91 [2.09, 4.09] | 3.912e-10 |
| **Smoker - Yes** | 1.31 [0.94, 1.84] | 0.114 | 1.27 [0.90, 1.78] | 0.171 |
| **Years of Education \* Age** |  |  | 1.005 [ 1.002, 1.009] | 0.003 |
| **N** | **878** |  |  |  |
| **Significant p-value** | **p < 0.007** |  | **p < 0.006** |  |
| **AIC** | **1108** |  | **1099** |  |
| **Pseudo-R<sup>2</sup> (Cragg-Uhler)** | **0.15** |  | **0.17** |  |

### Gender, Alcohol, Smoking, and Diabetes

Males have three times the odds of active TB than females (OR = 2.85, p < 0.001; Table 1 and Fig. 4). All logistic regression models showed insufficient statistical evidence for smoking (common risk factor model: OR =1.31, p = 0.11; Table 1, Table B in S1 Text), alcohol consumption (common risk factor model: OR = 1.05, p = 0.77; Table 1 and Table B in S1 Text) and diabetes (common risk factor model: OR =1.36, p =0.32; Table 1 and Table B in S1 Text) on TB risk. Despite the lack of significance, we note that smoking had an effect size in the expected direction (Fig. 4).

**Figure 4.**
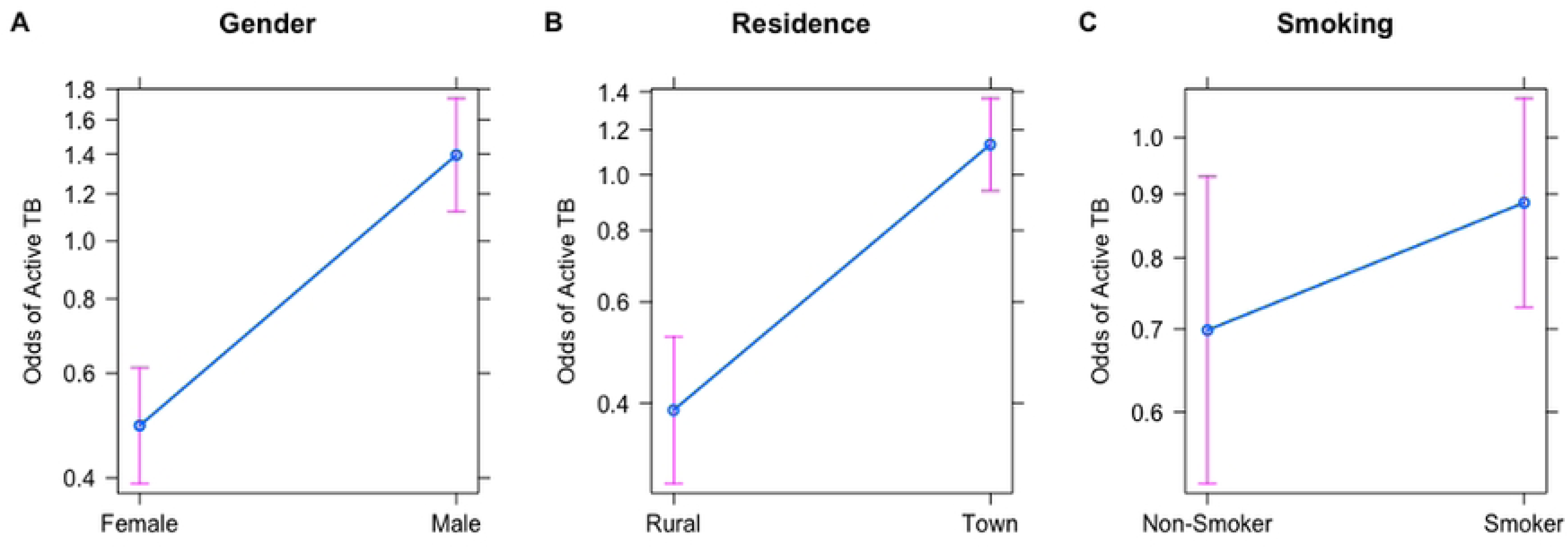
Effect Plots demonstrating the relationship between Active TB Status and A) Gender, B) Current Residence and C) Smoking. These plots are reported from the best-performing logistic regression model (SES model). Y-axes for all panels show the odds of active TB. We find that the odds of active TB are 3 times higher in Males. Individuals currently residing in Towns have about 2.5 times higher odds of active TB as compared to individuals currently residing in rural areas. Smoking slightly increases the odds of active TB but is not statistically significant.

### Age Interacts with SES

In the common risk factor model, age (OR= 0.996 [0.99, 1.00], p=0.55) and SES (OR = 0.947, p=0.0324; see Table 1) have no effect on TB risk. To examine this unexpected finding, we interacted age with years of education (proxy for SES). SES significantly affects TB status depending on age group (OR =1.005, p = 0.004, Table 1). The effect takes on a U-shaped relationship across ages, such that higher SES at younger ages (18-39 years old) is protective against TB, and higher SES at older ages (>59 years) increases risk (Fig. 5). Middle-aged individuals (40-59 years old) show no relationship between age and SES on TB risk (Fig. 5).

**Fig 5.**
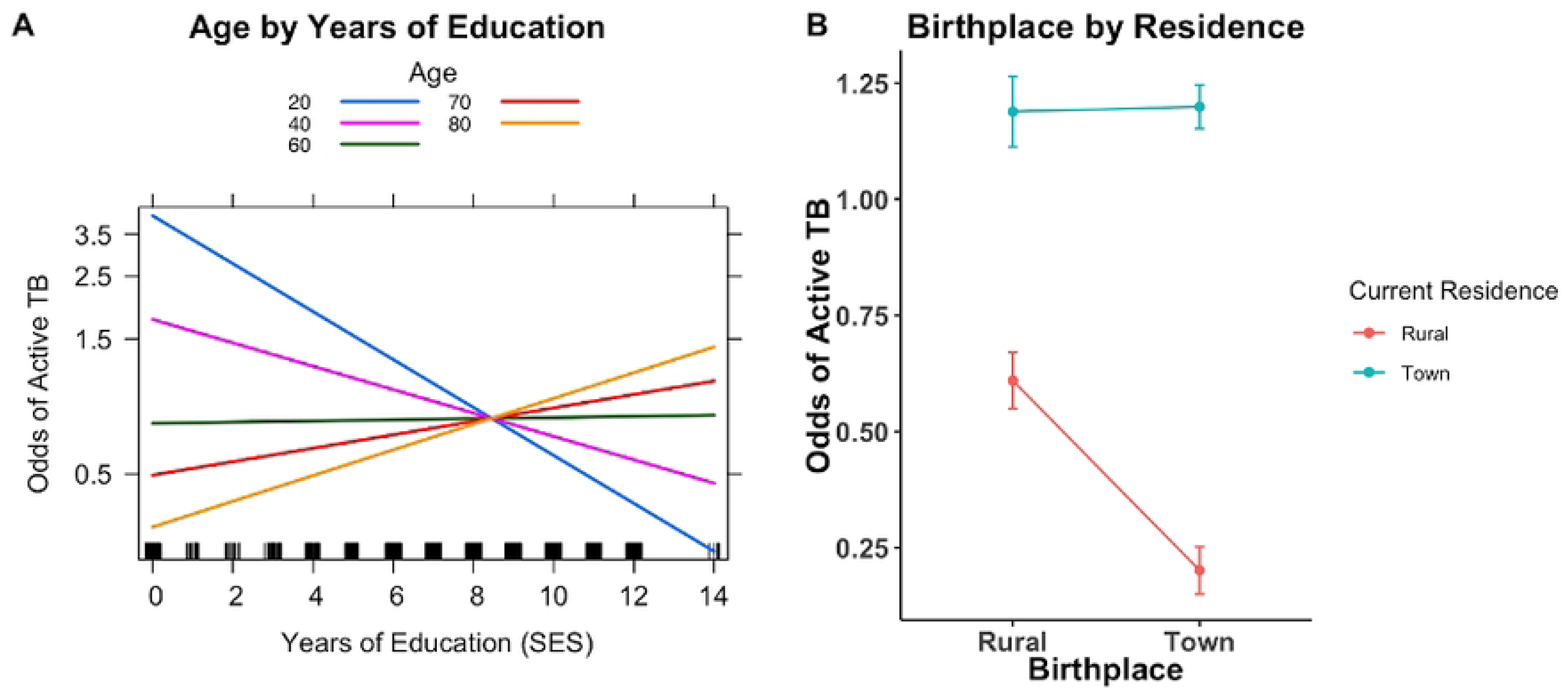
Logistic regression interaction plots. A) The odds of active TB by education level vary across age groups (shown above by the different color lines). More years of education decreases the odds of active TB in younger age groups, but this pattern reverses in the oldest age groups. In middle-aged individuals, there is no relationship between age and years of education. B) Effect plot from the residence model visualizing an interaction term between birthplace residence and current residence. Regardless of birthplace, the odds of active TB is highest in individuals who currently reside in towns. Individuals born in towns and currently residing in rural areas have the lowest odds of active TB.

### TB Risk is Highest in Towns

The odds of active TB were significantly higher for people residing in towns (common risk factor model: OR = 2.88 [2.07-4.03], p <0.0001; Table 1 and Fig. 4). For the residence model, we analyzed the impact of moving between rural areas and towns during an individual’s lifetime (birthplace by residence) on TB status. We expected to see a difference in odds for TB risk between life-long residents and those who have moved between locales. Under such a model, lifelong rural dwellers would have the lowest odds and lifelong town dwellers would have the highest odds. We set an interaction term between current residence and birthplace classified into town/rural; this interaction was marginally significant (OR = 3.05, p = 0.016; Table B in S1 Text). Our results show that regardless of birthplace, current residence in a town area increases the risk of active TB (Fig. 3B). Interestingly, individuals who were born in a town and later moved to rural areas are even more protected than individuals born and currently residing in rural areas (Fig. 3B).

### Random Forest Modeling

Similar to logistic regression, random forest is a binary classifier yet differs in that is robust against non-linear associations and unknown interactions (34). Random forest utilizes a permutation-based approach to generate a hierarchical list of important variables but is unable to quantify the “significance” between an independent and dependent variable.

5000 subsets of our dataset were used to grow 5000 classification trees using baseline variables as predictors for active TB status. The model assigned gender, current residence, and SES respectively as the overall top important independent variables (Fig II-A in S1 Text). Age, diabetes, alcohol, and smoking were classified as uninformative predictors for TB. The random forest model also stratified the variable importance by cases and controls. Gender was the top predictor for case status, followed by current residence and SES (Fig II-B in S1 Text). SES was the top predictor for control status followed by gender and current residence (Fig II-C in S1 Text). Interestingly, age had some predictive relevance for case status but was the worst-performing predictor for controls (Fig II-C in S1 Text). The model had an overall “out-of-bag” misclassification rate of 38%, with misclassification lower in controls (controls =30%, cases 48%; Supplementary Materials in S1 Text).

## Discussion

This present work represents the largest TB epidemiological study on a Northern Cape clinical population (n=878). In this study, we demonstrate the utility of population-based controls when disease exposure is known and transmission is community-spread (48) as seen in other studies in low-resource, high-burdened countries (9,49). Logistic regression and random forest models both show gender and residence as significant and important TB risk predictors. Random forest assigned SES as an important variable, and SES was only significant when interacting with age in logistic regressions. Neither smoking, alcohol consumption, nor diabetes is associated with increased TB risk in any model. Two logistic regression models, interacting SES by age (SES model), and birthplace by residence (residence model), had similar explanatory power, improving on the common risk factor model. This study demonstrates a possible unique historical context to South Africa, (post-)Apartheid differential effects between sociodemographic and health outcomes.

Age and TB risk have a general inverted U-shape relationship. During childhood, infants are at the greatest risk of TB decreasing through adolescence, increasing between 25-35 years old followed by a decrease, and another peak after 65 years (50,51). In our study population (≥18 years), the empirical odds of active TB increase with age and peak in the 49-58-year-old age group, followed by a steady decline in empirical odds after age 58 until the oldest age group (Fig. 2B). This drop in empirical odds after age 58 is most likely due to the mortality of individuals with TB, potentially a signal of survivor bias (52). This interpretation is seen in the shifting proportions of cases and controls across age groups (Fig. 2A). From ages 21 - 58, most of the population are cases, and from ages 59 - 88 most of the population controls (Fig. 2A).

Age was neither a significant (logistic regression) nor an important (random forest) variable except when interacted with SES. SES’s protective effect on TB risk is most prominent among 18-39 year-olds and becomes a risk factor among the eldest individuals (>65 years; Fig. 5A)—those who grew up and reached adulthood during Apartheid (Fig. 5A). Higher SES increasing TB risk is contrary to findings in populations in the United States and Mexico (51).

This unique pattern may reflect South Africa’s recent history of Apartheid and post-Apartheid societal and economic shifts. During Apartheid, individuals from historically marginalized backgrounds had limited career options, but some were able to become teachers, police officers, or nurses. Such occupations are associated with higher education requirements and would have facilitated access to larger salaries, transportation, and mobility.

Higher SES could result in apparent greater odds of TB because these individuals would have had better access to healthcare both facilitating diagnosis and treatment. Universal access to education increased post-Apartheid but given the wide variation of years of education among the youngest generations, it likely still covaries with SES. Given the unusual interaction here between age and years of education, future work should validate additional SES measures to resolve mechanisms of TB risk.

Consistent with previous research (53–56), we find TB risk is associated with living in larger towns. In our prior work, mobility in the Northern and Western Cape populations changed over the past 3 generations with the highest levels of mobility in the grandparental generation (57). Therefore, we tested whether mobility (different birthplace and residence) affected TB risk. As expected, the protective effect of living rurally vanishes when someone moves to a larger town. Further, the individuals with the lowest TB risk are those born in a town and move to a rural area. These findings are consistent with TB exposure nearing ∼ 90% by 25-30 years old (13), with transmission occurring via community contacts during adolescence and adulthood. We hypothesize that those born in towns who later moved to a rural area benefit from both BCG vaccination and decreased adult exposure thereby overall decreasing their odds of TB. BCG vaccination is standard for children in South Africa, however, children in rural areas may have lower vaccination rates (observation communicated by clinical staff in the study catchment). Future work should consider collecting birthplace in addition to current residence to better identify TB risk.

Invariably across studies, men are on average 1.7 times more likely to have TB (58–60). Sex biases like this are common in other infectious diseases (61,62) and are attributable to an intersection of sex (biological factors, e.g., immune function) and gender (social and behavioral factors, e.g., risk-taking behavior) (63). Despite smoking not being a significant TB risk, we found 75.5% of men smoke compared to 55.8% of women, indicating at least some gender differences in risky behaviors in the Northern Cape population.

Smoking and alcohol consumption has been shown to increase TB risk and mortality in the Northern Cape and at the national level (64–67). In our models smoking had the expected effect on TB risk and alcohol consumption had no effect. Both variables lacked statistical power in regression models and failed to meet any level of importance in the random forest model. Self-reporting biases in observational studies like this one are a concern for variables like smoking, alcohol consumption, and SES measures (68). Our sample, however, reports much higher levels of smoking compared to large-scale national surveys (e.g., (69), men: 75.5.% vs. 41%; women: 55.8% vs. 21%, respectively suggesting minimal self-report bias in our study. It is possible that these weak effects of smoking and alcohol observed from our models are due to our method of binary classification. We collected fine-scale smoking and alcohol phenotypes (Supplementary Methods in S1 Text) but because of the high missingness of these phenotypes, we ultimately classified participants as Smokers/Non-smokers and Drinkers/Non-Drinkers. This stratification may mask the heterogeneity of drinking and smoking behaviors such as casual and binge substance use or differences in the types of alcohol and smoking materials consumed. Further TB epidemiological studies in the Northern Cape should explore these smoking and alcohol phenotypes in more detail.

Active TB progression is a multifactorial process involving the environment, genetics, and their interaction (1,4). Our results from the NCTB cohort indicate that sociodemographic variables strongly impact active TB risk. Effects that are unique to the Northern Cape Province may reflect how changes in the pre- to post-apartheid environment modified social factors, such as SES and mobility, which in turn impacted lifetime TB risk. This work provides a baseline to design well-informed future studies, such as exploring host genetic correlates of active TB progression in this population (Supplementary Discussion in S1 Text).

## Data Availability

All R scripts for statistical data analyses and data visualization are available at https://github.com/oshiomah1/NCTB-Epidemiology-Project. The relevant raw data will be deposited in EGA database. https://github.com/oshiomah1/NCTB-Epidemiology-Project/tree/main

## Acknowledgments

We would like to thank all the participant communities in the Northern Cape for their continued trust and support in helping us undertake this project. We would also like to thank our community research assistants and translators who assisted in data collection for the project. We are grateful to Prof Faadiel Essop, Dr. Desiree Petersen, Prof Eileen Hoal, and Prof Leslie Swartz for a close reading of this manuscript. Finally, we want to thank the Department of Health in the Northern Cape Province, South Africa for their continued support of the project.

## S1 Text

Supplementary Methods

Supplementary Results

Supplementary Discussion

**Table A.** Descriptive Statistics of study variables, stratified by case/control status

**Table B.** Odds ratios and p-values for the Demographic and Socio Behavioral Variables used in the Residence Model

**Table C.** Descriptive statistics of Raw Dataset (Before imputation and implementing HIV exclusion criteria

**Table D:** Missingness and Imputation Metrics

**Fig I. Correlations Among Demographic and Medical Variables for Entire Cohort**. Pearson correlation coefficients were calculated for select demographic, behavioral, and medical covariates in our dataset of 878 individuals. Correlations with significant p-values (p<0.05) are denoted with an asterisk.

**Fig II. Hierarchical list of important variables from Random Forest Model in A) All individuals (n= 878) B) Cases (n =374) and `C) Controls (n =504).`** Random subsets of all 7 variables on the y-axis were used to grow 5000 trees to classify participants into cases and controls. Mean decrease accuracy was computed by the differences in classification error between the “out-of-bag” dataset and a randomly permuted sample (*Supplementary methods in S1 text)*. Variables with higher mean decrease accuracy are most important for case-control classification. Predictor variables with mean decrease accuracy values close to zero have no effect on classification while negative values worsen the ability of the model to classify TB status.

## References

1. WHO. Global Tuberculosis Report 2021 [Internet]. 2021 Oct [cited 2022 Aug 8]. Available from: https://www.who.int/publications/i/item/9789240037021

2. Brites D, Gagneux S. Co-evolution of Mycobacterium tuberculosis and Homo sapiens. Immunol Rev. 2015 Mar;264(1):6–24.

3. Sharma SK, Mohan A. Extrapulmonary Tuberculosis. Indian J Med Res [Internet]. 2004 [cited 2022 Nov 11];120(4)(316). Available from: https://www.proquest.com/openview/330577dc52a107765d6adb9b1168c6e6/1?pq-origsite=gscholar&cbl=37533

4. Glaziou P, Falzon D, Floyd K, Raviglione M. Global epidemiology of tuberculosis. Semin Respir Crit Care Med. 2013;34(1):3–16.

5. Lacerda SNB, De Abreu Temoteo RC, De Figueiredo TMRM, De Luna FDT, De Sousa MAN, De Abreu LC, et al. Individual and social vulnerabilities upon acquiring tuberculosis: A literature systematic review. Int Arch Med. 2014 Jul;7(1):1–8.

6. Fok A, Numata Y, Schulzer M, FitzGerald MJ. Risk factors for clustering of tuberculosis cases: a systematic review of population-based molecular epidemiology studies [Review Article]. Int J Tuberc Lung Dis. 2008 May 1;12(5):480–92.

7. Statistics South Africa. Mortality and Causes of death in South Africa: Findings from death notification 2018 [Internet]. 2018 [cited 2022 Apr 4]. Available from: https://www.statssa.gov.za/publications/P03093/P030932017.pdf

8. South African National Department of Health, South African Medical Research Council;, Human Sciences Research Council, National Institute for Communicable Diseases;, World Health Organization, United States Agency for International Development, et al. The First National TB Prevalence Survey | South Africa 2018 [Internet]. 2018 [cited 2021 Sep 11]. Available from: https://www.google.com/search?client=safari&rls=en&q=the+first+national+tb+prevalence+survey+south+africa+2018&ie=UTF-8&oe=UTF-8

9. Duchen D, Vergara C, Thio CL, Kundu P, Chatterjee N, Thomas DL, et al. Pathogen exposure misclassification can bias association signals in GWAS of infectious diseases when using population-based common control subjects. Am J Hum Genet. 2023 Feb;110(2):336– 48.

10. Verver S, Warren RM, Munch Z, Richardson M, al et. Proportion of tuberculosis transmission that takes place in households in a high-incidence area. The Lancet. 2004 Jan 17;363(9404):212–4.

11. Middelkoop K, Mathema B, Myer L, Shashkina E, Whitelaw A, Kaplan G, et al. Transmission of Tuberculosis in a South African Community With a High Prevalence of HIV Infection. J Infect Dis. 2015 Jan 1;211(1):53–61.

12. Gallant CJ, Cobat A, Simkin L, Black GF, Stanley K, Hughes J, et al. Impact of age and sex on mycobacterial immunity in an area of high tuberculosis incidence. Int J Tuberc Lung Dis. 2010 Aug;14(8):952–9.

13. Bunyasi EW, Schmidt BM, Abdullahi LH, Mulenga H, Tameris M, Luabeya A, et al. Prevalence of latent TB infection and TB disease among adolescents in high TB burden countries in Africa: a systematic review protocol. BMJ Open. 2017 Mar 1;7(3):e014609.

14. Wood R, Liang H, Wu H, Middelkoop K, Oni T, Rangaka MX, et al. Changing prevalence of tuberculosis infection with increasing age in high-burden townships in South Africa. Int J Tuberc Lung Dis Off J Int Union Tuberc Lung Dis. 2010 Apr;14(4):406–12.

15. Uys P, Brand H, Warren R, Spuy G van der, Hoal EG, Helden PD van. The Risk of Tuberculosis Reinfection Soon after Cure of a First Disease Episode Is Extremely High in a Hyperendemic Community. PLOS ONE. 2015 Dec 9;10(12):e0144487.

16. Karlsson EK, Kwiatkowski DP, Sabeti PC. Natural selection and infectious disease in human populations. Nat Rev Genet. 2014 Apr;15(6):379–93.

17. Möller M, Kinnear CJ, Orlova M, Kroon EE, van Helden PD, Schurr E, et al. Genetic Resistance to Mycobacterium tuberculosis Infection and Disease. Front Immunol [Internet]. 2018 [cited 2022 Nov 11];9. Available from: https://www.frontiersin.org/articles/10.3389/fimmu.2018.02219

18. Uren C, Hoal EG, Möller M. Mycobacterium tuberculosis complex and human coadaptation: a two-way street complicating host susceptibility to TB. Hum Mol Genet. 2021 Apr 26;30(R1):R146–53.

19. Leal DF, da VB, Silva MNS da Fernandes DCR de O, Rodrigues JCG, Barros MC da C, Pinto PD do C, et al. Amerindian genetic ancestry as a risk factor for tuberculosis in an amazonian population. PLOS ONE. 2020 Jul 16;15(7):e0236033.

20. Luo Y, Suliman S, Asgari S, Amariuta T, Baglaenko Y, Martínez-Bonet M, et al. Early progression to active tuberculosis is a highly heritable trait driven by 3q23 in Peruvians. Nat Commun. 2019;10(1):1–10.

21. Asgari S, Luo Y, Huang CC, Zhang Z, Calderon R, Jimenez J, et al. Higher native Peruvian genetic ancestry proportion is associated with tuberculosis progression risk. Cell Genomics. 2022 Jul 13;2(7):100151.

22. Chimusa ER, Zaitlen N, Daya M, Möller M, Helden PD van, Nicola JM, et al. Genome-wide association study of ancestry-specific TB risk in the South African coloured population. Hum Mol Genet. 2014;23(3):796–809.

23. Daya M, van der Merwe L, van Helden PD, Möller M, Hoal EG. The role of ancestry in TB susceptibility of an admixed South African population. Tuberculosis. 2014 Jul 1;94(4):413– 20.

24. Nava-Aguilera E, Andersson N, Harris E, Mitchell S, Hamel C, Shea B, et al. Risk factors associated with recent transmission of tuberculosis: systematic review and meta-analysis. Int J Tuberc Lung Dis Off J Int Union Tuberc Lung Dis. 2009 Jan;13(1):17–26.

25. Bates MN, Khalakdina A, Pai M, Chang L, Lessa F, Smith KR. Risk of tuberculosis from exposure to tobacco smoke: A systematic review and meta-analysis. Arch Intern Med. 2007 Feb;167(4):335–42.

26. Rehm J, Samokhvalov AV, Neuman MG, Room R, Parry C, Lönnroth K, et al. The association between alcohol use, alcohol use disorders and tuberculosis (TB). A systematic review. BMC Public Health. 2009 Dec;9(1):450.

27. Fiske CT, Hamilton CD, Stout JE. Alcohol use and clinical manifestations of tuberculosis. J Infect. 2009 May;58(5):395–401.

28. Imtiaz S, Shield KD, Roerecke M, Samokhvalov AV, Lönnroth K, Rehm J. Alcohol consumption as a risk factor for tuberculosis: meta-analyses and burden of disease. Eur Respir J. 2017 Jul;50(1):1700216.

29. Barnes PF. Weighing Gold or Counting Spots. Am J Respir Crit Care Med. 2006 Oct;174(7):731–2.

30. Stout JE, Menzies D. Predicting Tuberculosis Does the IGRA Tell the Tale? Am J Respir Crit Care Med. 2008 May 15;177(10):1055–7.

31. Kroon EE, Kinnear CJ, Orlova M, Fischinger S, Shin S, Boolay S, et al. An observational study identifying highly tuberculosis-exposed, HIV-1-positive but persistently TB, tuberculin and IGRA negative persons with M. tuberculosis specific antibodies in Cape Town, South Africa. EBioMedicine. 2020 Nov 1;61:103053.

32. Verrall AJ, Netea MG, Alisjahbana B, Hill PC, van Crevel R. Early clearance of Mycobacterium tuberculosis: a new frontier in prevention. Immunology. 2014 Apr;141(4):506–13.

33. Smith MH, Myrick JW, Oyageshio O, Uren C, Saayman J, Boolay S, et al. Epidemiological correlates of overweight and obesity in the Northern Cape Province, South Africa. PeerJ. 2023 Feb 9;11:e14723.

34. Bi Q, Goodman KE, Kaminsky J, Lessler J. What is Machine Learning? A Primer for the Epidemiologist. Am J Epidemiol. 2019 Oct 21;kwz189.

35. Iwendi C, Bashir AK, Peshkar A, Sujatha R, Chatterjee JM, Pasupuleti S, et al. COVID-19 Patient Health Prediction Using Boosted Random Forest Algorithm. Front Public Health [Internet]. 2020 [cited 2022 Aug 29];8. Available from: https://www.frontiersin.org/articles/10.3389/fpubh.2020.00357

36. Loef B, Wong A, Janssen NAH, Strak M, Hoekstra J, Picavet HSJ, et al. Using random forest to identify longitudinal predictors of health in a 30-year cohort study. Sci Rep. 2022 Jun 20;12(1):10372.

37. Ooka T, Johno H, Nakamoto K, Yoda Y, Yokomichi H, Yamagata Z. Random forest approach for determining risk prediction and predictive factors of type 2 diabetes: large-scale health check-up data in Japan. BMJ Nutr Prev Health [Internet]. 2021 Jun 1 [cited 2022 Aug 29];4(1). Available from: https://nutrition.bmj.com/content/4/1/140

38. 38. Bolker B, Warnes G, Lumley T. gtools: Various R Programming Tools. R package version 3.9.3 [Internet]. 2022. Available from: https://CRAN.R-project.org/package=gtools}

39. Fox J. Effect Displays in R for Generalised Linear Models. J Stat Softw. 2003 Jul 22;8:1–27.

40. Baker PA. From Apartheid to Neoliberalism: Health Equity in Post-Apartheid South Africa. Int J Health Serv. 2010 Jan 1;40(1):79–95.

41. Maphumulo WT, Bhengu BR. Challenges of quality improvement in the healthcare of South Africa post-apartheid: A critical review. Curationis. 2019 May 29;42(1):e1–9.

42. Mhlanga D, Garidzirai R. The Influence of Racial Differences in the Demand for Healthcare in South Africa: A Case of Public Healthcare. Int J Environ Res Public Health. 2020 Jan;17(14):5043.

43. Alexander DH, Novembre J, Lange K. Fast model-based estimation of ancestry in unrelated individuals. Genome Res. 2009 Sep;19(9):1655.

44. Behr AA, Liu KZ, Liu-Fang G, Nakka P, Ramachandran S. pong: fast analysis and visualization of latent clusters in population genetic data. Bioinforma Oxf Engl. 2016 Sep 15;32(18):2817–23.

45. Statistics South Africa. Provincial profile: Northern Cape Community Survey 2016, Report 03-01-14. Private Bag X44, Pretoria, 0001; 2018.

46. Adhikari M. The Sons of Ham: Slavery and the Making of Coloured Identity. South Afr Hist J. 1992 Nov 1;27(1):95–112.

47. LaValley MP. Logistic Regression. Circulation. 2008 May 6;117(18):2395–9.

48. Munch Z, Van Lill SWP, Booysen CN, Zietsman HL, Enarson DA, Beyers N. Tuberculosis transmission patterns in a high-incidence area: a spatial analysis. Int J Tuberc Lung Dis Off J Int Union Tuberc Lung Dis. 2003 Mar;7(3):271–7.

49. Lienhardt C, Fielding K, Sillah J, Bah B, Gustafson P, Warndorff D, et al. Investigation of the risk factors for tuberculosis: a case–control study in three countries in West Africa. Int J Epidemiol. 2005 Aug 1;34(4):914–23.

50. Davies PDO. Risk factors for tuberculosis. Monaldi Arch Chest Dis. 2005 Mar;63(1):37–46.

51. Scordo JM, Aguillón-Durán GP, Ayala D, Quirino-Cerrillo AP, Rodríguez-Reyna E, Mora-Guzmán F, et al. A prospective cross-sectional study of tuberculosis in elderly Hispanics reveals that BCG vaccination at birth is protective whereas diabetes is not a risk factor. PLOS ONE. 2021 Jul;16(7):e0255194–e0255194.

52. Swanson DM, Anderson CD, Betensky RA. Hypothesis Tests for Neyman’s Bias in Case-Control Studies. J Appl Stat. 2018;45(11):1956–77.

53. Beiranvand R, Karimi A, Delpisheh A, Sayehmiri K, Soleimani S, Ghalavandi S. Correlation Assessment of Climate and Geographic Distribution of Tuberculosis Using Geographical Information System (GIS). Iran J Public Health. 2016 Jan;45(1):86–93.

54. Hoffner S, Hadadi M, Rajaei E, Farnia P, Ahmadi M, Jaberansari Z, et al. Geographic characterization of the tuberculosis epidemiology in iran using a geographical information system. Biomed Biotechnol Res J BBRJ. 2018;2(3):213.

55. Ncayiyana JR, Bassett J, West N, Westreich D, Musenge E, Emch M, et al. Prevalence of latent tuberculosis infection and predictive factors in an urban informal settlement in Johannesburg, South Africa: a cross-sectional study. BMC Infect Dis. 2016 Nov 8;,16(1):661.

56. Sikalengo G, Hella J, Mhimbira F, Rutaihwa LK, Bani F, Ndege R, et al. Distinct clinical characteristics and helminth co-infections in adult tuberculosis patients from urban compared to rural Tanzania. Infect Dis Poverty. 2018 Mar 24;7(1):24.

57. Reynolds A, Grote MN, Myrick JW, Al-Hindi DR, Siford RL, Mastoras M, et al. Persistence of matrilocal post-marital residence across multiple generations in Southern Africa [Internet]. SocArXiv; 2022 [cited 2022 Nov 11]. Available from: https://osf.io/preprints/socarxiv/7qfns/

58. Hertz D, Schneider B. Sex differences in tuberculosis. Semin Immunopathol. 2019 Mar;41(2):225–37.

59. Holmes CB, Hausler H, Nunn P. A review of sex differences in the epidemiology of tuberculosis. Int J Tuberc Lung Dis. 1998 Feb 1;2(2):96–104.

60. Neyrolles O, Quintana-Murci L. Sexual Inequality in Tuberculosis. PLOS Med. 2009 Dec 22;6(12):e1000199.

61. WHO (Western Pacific Region). Taking sex and gender into account in emerging infectious disease programme: an analytical framework [Internet]. 2007 [cited 2022 Aug 29]. Available from: https://www.who.int/publications-detail-redirect/9789290615323

62. Wizemann TM, Pardue ML. Exploring the Biological Contributions to Human Health [Internet]. National Academies Press (US); 2001 [cited 2022 Aug 29]. Available from: https://www.ncbi.nlm.nih.gov/books/NBK222288/

63. Lawry LL, Lugo-Robles R, McIver V. Improvements to a framework for gender and emerging infectious diseases. Bull World Health Organ. 2021 Sep 1;99(9):682–4.

64. Harling G, Ehrlich R, Myer L. The social epidemiology of tuberculosis in South Africa: A multilevel analysis. Soc Sci Med. 2008;66(2):492–505.

65. Peltzer K, Louw J, Mchunu G, Naidoo P, Matseke G, Tutshana B. Hazardous and Harmful Alcohol Use and Associated Factors in Tuberculosis Public Primary Care Patients in South Africa. Int J Environ Res Public Health. 2012 Sep;9(9):3245–57.

66. Sitas F, Urban M, Bradshaw D, Kielkowski D, Bah S, Peto R. Tobacco attributable deaths in South Africa. Tob Control. 2004 Dec;13(4):396–9.

67. Wessels J, Walsh CM, Nel M. Smoking habits and alcohol use of patients with tuberculosis at Standerton Tuberculosis Specialised Hospital, Mpumalanga, South Africa. Health SA SA Gesondheid. 2019 Oct 8;24:1146.

68. Althubaiti A. Information bias in health research: definition, pitfalls, and adjustment methods. J Multidiscip Healthc. 2016 May 4;9:211–7.

69. National Department of Health (NDoH), Statistics South Africa, South African Medical Research Coucil, ICF. South Africa Demographic and Health Survey 2016. [Internet]. [cited 2022 Aug 8]. Available from: https://dhsprogram.com/pubs/pdf/FR337/FR337.pdf

